# Ventricular anatomical complexity and gender differences impact predictions from computational models

**DOI:** 10.1101/2022.01.28.22269815

**Authors:** Pablo González-Martín, Federica Sacco, Constantine Butakoff, Rubén Doste, Carlos Bederián, Lilian K Gutierrez Espinosa de los Monteros, Guillaume Houzeaux, Paul A. Iaizzo, Tinen L. Iles, Mariano Vázquez, Jazmin Aguado-Sierra

**Affiliations:** Barcelona Supercomputing Center, Barcelona, Spain; ELEM Biotech S.L., Barcelona, Spain; Physense, Department of Information and Communication Technologies, Universitat Pompeu Fabra, Barcelona, Spain; Department of Computer Science, University of Oxford, Oxford, UK; Instituto de Física Enrique Gaviola - CONICET, Córdoba, Argentina; Centro Nacional de Investigaciones Cardiovasculares Carlos III, Madrid, Spain; Visible Heart^®^ Laboratories, Department of Surgery and the Institute for Engineering in Medicine, University of Minnesota, Minneapolis, USA; University of Minnesota Medical School, Minneapolis, USA

**Author notes:** These authors contributed equally to this work (Pablo González-Martín, Federica Sacco).

## Abstract

The aim of this work was to analyze the influence of the gender and anatomical details (trabeculations and false tendons) on the electrophysiology of healthy and infarcted myocardium and their relation to the ventricular tachycardia (VT) inducibility.

To this end, four anatomically normal, human, biventricular geometries (two male, two female), with identifiable trabeculations, were obtained from high-resolution, ex-vivo MRI and represented by detailed and smoothed geometrical models (with and without the trabeculations). Additionally one model was augmented by a scar. The electrophysiology finite element model (FEM) simulations were carried out, using O’Hara-Rudy human myocyte model, and the comparison of detailed vs smooth, male vs female, as well as normal vs infarcted anatomy was carried out. Finally, the infarcted case was subjected to standard clinical programmed stimulus S1-S4 protocol to identify geometry- and gender-specific VT inducibility.

All female hearts presented action potential prolongation and QRS interval lengthening. Smooth geometries showed different QRS pattern and T wave magnitude in comparison to the corresponding trabeculated geometries. A variety of sustained VTs were obtained in the detailed and smoothed male geometries at different pacing locations, which provide evidence of the geometry-dependent differences regarding the prediction of the locations of reentry channels. In the female phenotype, sustained VTs were induced in both detailed and smooth geometries with RV apex pacing, however no consistent reentry channels were identified.

Anatomical and physiological cardiac features play an important role defining risk in cardiac disease. These are often excluded from electrophysiological simulations. The assumption that the cardiac endocardium is smooth may inaccurately predict different reentry channel locations in tachycardia inducibility studies.

## Introduction

Imaging-based cardiac computational simulations have opened a new paradigm for personalized medicine applied to complex structural pathologies in search for a better understanding and stratification of disease. Computational tools are also being recognized as key contributors to the reduction of the time and economical burden of regulatory pathways for both the pharmaceutical and the medical device industry aiming towards a more efficient design and development of drugs, treatments, and ultimately to achieve better outcomes in the clinic.

Cardiac electrophysiology modelling has been a primordial target for computer simulations. Seventy years of research on mathematical modelling of cardiac electrophysiology has produced a wealth of information regarding single cell dynamics that led to various mathematical models describing the ventricular function of the human heart [1]. However, even when detailed mathematical cell models are available, the geometry of the ventricle, used for the simulations is often simplified. The simplifications are dictated by the imaging modality available for the model construction and computational power available. The most typical imaging modality used for model construction is Magnetic Resonance Imaging (MRI), acquired in slices, with, typically, good within-slice resolution but 5-10mm distance between slices. This low spatial resolution is insufficient for capturing the complexity of human endocardium (i.e., trabeculae and false tendons). It is therefore important to quantify the potential errors produced by the geometrical simplifications, given the imaging data obtained in the every-day clinic, as the endocardial structures may create electrical bridges and may play a crucial role in efficient action potential propagation.

Functional gender differences is another aspect of the computational cardiac models that is generally ignored. Understanding the gender-specific cardiac pathophysiology is still in its infancy. These differences may play an important role in arrhythmogenesis [2]. For example, Yang et al. [3], reported genomic-based differences within ion channel expressions, leading to longer cardiac action potential durations (APDs) in females than in males. Female human hearts have been shown to elicit lower expressions of the genes responsible for cardiac repolarizing potassium currents, and Connexin43. Considering the differences in ion channel sub-unit expressions is important when studying gender-specific physio-pathologies, in particular gender-specific arrhythmic risks.

All human cardiac ventricles have a sponge-like architecture inside the cavity, different in every heart [4]. False tendons establish fast conducting shortcuts within the ventricular cavities, but their small diameter makes them impossible to reproduce on patient-specific cardiac computational models using conventional clinical imaging tools. Nevertheless, they play a role in the overall cardiac pathophysiology.

To date, there have been a few computational models studying the influences of the position and amount of both trabeculae and FTs on the overall cardiac electrophysiology (Bishop et al. [5], Bordas et al. [6], Lange et al. [7], Connolly et al. [8] and Galappaththinge et al. [9]). Nevertheless, none of these previous works explore their effect on ventricular tachycardia simulations and the reentry channel locations as related to gender-specific risk prediction.

The primary aim of our work was to analyze the influences of both highly detailed anatomical endocardial structures and gender phenotypes on human electrophysiology. To do so, we studied four biventricular, anatomically normal human heart models, reconstructed from high-resolution *ex-vivo* MRI data: one male adult, one male child and two female adults and their smoothed equivalents. The relative influences of gender phenotypes and endocardial sub-structures were studied by applying both phenotypes to all detailed and smoothed geometries and calculating their pseudo-ECG. In this way we could critically analyze the influences of gender on associated cardiac electrophysiology as well as quantify the errors (root mean square errors) on the pseudo-ecg introduced computationally by neglecting endocardial sub-structures such as trabeculae and false tendons. Further, a full clinical programmed stimulation protocol to assess ventricular tachycardia inducibility was performed on one of these cardiac models after a myocardial infarction event was introduced: i.e., into both the detailed and the smoothed geometries. Myocardial infarction information was obtained from a gadolinium-enhanced MRI clinical imaging protocol performed on a different clinical patient.

## Materials and methods

### Human Anatomical Data

Four biventricular models, two males (one child and one adult) and two females, were segmented from high-resolution MRI data of *ex-vivo* perfusion fixed human hearts. These hearts were obtained from organ donors whose hearts were deemed not viable for transplant throught the local procurement organization, LifeSource. These specimens are part of the Visible Heart® Laboratories human heart library at the University of Minnesota. The uses of these heart specimens for research were given the appropriate written consent, witnessed by LifeSource, from the donor and their families, the University of Minnesota’s Institution Review Board and LifeSource Research Committee. All data has been anonymized from its source. The hearts were cannulated and perfusion fixed under a pressure of approximately 40 − 50 mmHg with 10% phosphate buffered formalin in order to preserve the hearts in an end-diastolic volume. The images of the fixed hearts were then acquired via a 3T Siemens scanner with 0.44 *×* 0.44 mm in-plane resolution and slice thickness of 1 to 1.7 mm.

### Anatomical Mesh Construction

The four hearts utilized for the simulation were selected based on their average mid-cavity LV thickness in order to identify normal, healthy human heart values [10]: 0.6 ± 3 0.11 cm in male hearts and 0.53 ± 0.09 cm in the female hearts.

Corresponding smoothed models were then generated from the detailed ones from the manual delineation of the smoothed endocardial surfaces for each image slice, so to maintain the overall outline of the geometry. The four models and their corresponding smoothed geometries are shown in Fig 1. A detailed description of the methods employed to create the anatomical meshes can be found in the Supplemental Data section S 1.1.

**Fig 1.**
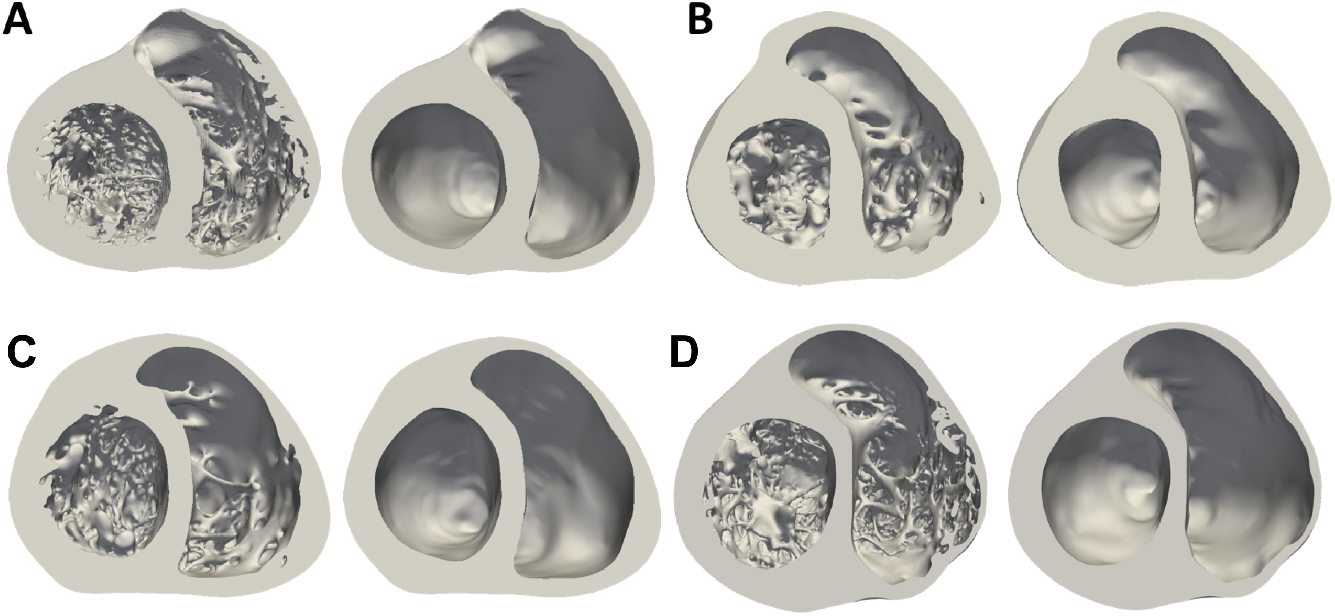
Male (**A**-**B**) and female (**C**-**D**) anatomically detailed and corresponding smoothed biventricular geometries. Mid-cavity section.

The myocardial volume of each biventricular heart (detailed and smoothed) anatomy is reported in Table 1, along with the volume occupied by the detailed endocardial structures (percentage of trabeculations), the number of long FTs (longer than 1 cm), the average LV thicknesses and tetrahedral-element-mesh information.

**Table 1.** Detailed and smoothed biventricular models description.

| Heart Geometry | A |  | B |  | C |  | D |  |
| --- | --- | --- | --- | --- | --- | --- | --- | --- |
|  | Detailed | Smoothed | Detailed | Smoothed | Detailed | Smoothed | Detailed | Smoothed |
| Myocardial Volume [$cm^3$] | 394.2 | 329.8 | 199.6 | 180.5 | 170.6 | 155.8 | 299.2 | 268 |
| Trabecular Volume [%] | 16.3 |  | 9.6 |  | 8.7 |  | 10.4 |  |
| N° of FTs $\geq 1cm$ | 3 | // | 0 | // | 4 | // | 3 | // |
| Average LV thickness [$cm$] | 0.85 | | 0.83 | | 0.58 | | 0.76 | |
| Volumetric Mesh Elements | 86,318,429 | 72,165,805 | 43,677,705 | 39,502,627 | 37,336,461 | 34,089,714 | 65,501,799 | 58,652,154 |
| Volumetric Mesh Points | 14,994,563 | 12,411,277 | 7,530,021 | 6,796,739 | 6,496,017 | 5,896,926 | 11,416,445 | 10,793,625 |

All volumetric meshes had a regular maximum element side length of 0.4 mm. Wireframe images of the four detailed tetrahedral meshes are shown in the Supplemental Data, Figure 1.

### Electrophysiological simulations

The monodomain approximation to electrical propagation was employed as described by Vazquez et al. [11] The reaction-diffusion system was solved using Alya, an in-house HPC code developed at the Barcelona Supercomputing Center. Briefly, the system to be solved is defined as:

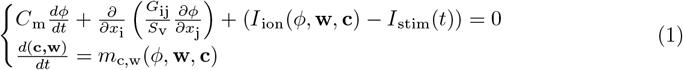

where *ϕ* is the membrane potential, *C*_*m*_ is the membrane capacitance, *G*_ij_ is the conductivity tensor, *S*_v_ is the surface to volume ratio of the cardiomyocyte, I_stim_(t) is the applied current (which initiates activation), I_ion_ is the ionic current term and **w** and **c** are the ionic currents gating variables and the intracellular concentrations, respectively. The O’Hara-Rudy [12] human cardiac ionic model with the modifications suggested in Dutta et al. [13] was employed to compute *I*_*ion*_.

In this work transmural myocyte heterogeneity was considered by assigning different electrophysiological and cellular properties to the following parts of the cardiac walls: endocardial (inner 30%), mid-myocardial (middle 40%) and epicardial (outer 30%) [12]. Apex-to-base cell heterogeneity was achieved by modifying the conductance of the slow delayed rectifier potassium current *IK*_*s*_ following a linear gradual decay from apex to base.

In order to study the influence of human gender phenotypes on cardiac electrophysiology, they were modelled incorporating into the O’Hara-Rudy model [12] all the changes to ion channels sub-unit expression that are listed in [3], which are reported in the Supplemental Data Table 1. Initial conditions for either male or female electrophysiology phenotype were calculated for a cardiac cycle duration of 600 ms.

Finally, a higher diffusion was assigned to a one-element layer on the endocardial surface in order to account for the fast conduction of the Purkinje fibers. The existence of this high conduction layer is important to simulate a physiological conduction velocity in the epicardium and to achieve a normal total activation time. Detailed analyses, described in Section 2.4, were carried out with the four models to define physiological diffusion values for both myocardium and endocardium in all anatomies.

All the electrophysiological simulations were carried out on MareNostrum 4 supercomputer, using the Barcelona Supercomputing Centre (BSC)’s in-house, parallel multi-physics, HPC solver Alya [14–16], with computing time provided by the Red Española de Supercomputacion, RES (Project BCV-2019-3-0009).

### Fiber Orientation Model

A rule-based model [17] was used to assign fibre orientations to all biventricular models. Specifically, fiber orientations within the anatomically detailed hearts were generated by automatically detecting trabeculations through the use of the magnitude of the transmural gradient (gradient was obtained by solving Laplace equation between the endocardium and the epicardium) and assigning them orientations along the longitudinal direction, following their centreline. The transition between cardiomyocytes within trabeculations and endocardium is smoothed according to the angle formed between the micro-structures and the endocardial wall. A close-up of the fibre orientation in both LV and RV of biventricular model **A** is shown in Fig 2.

**Fig 2.**
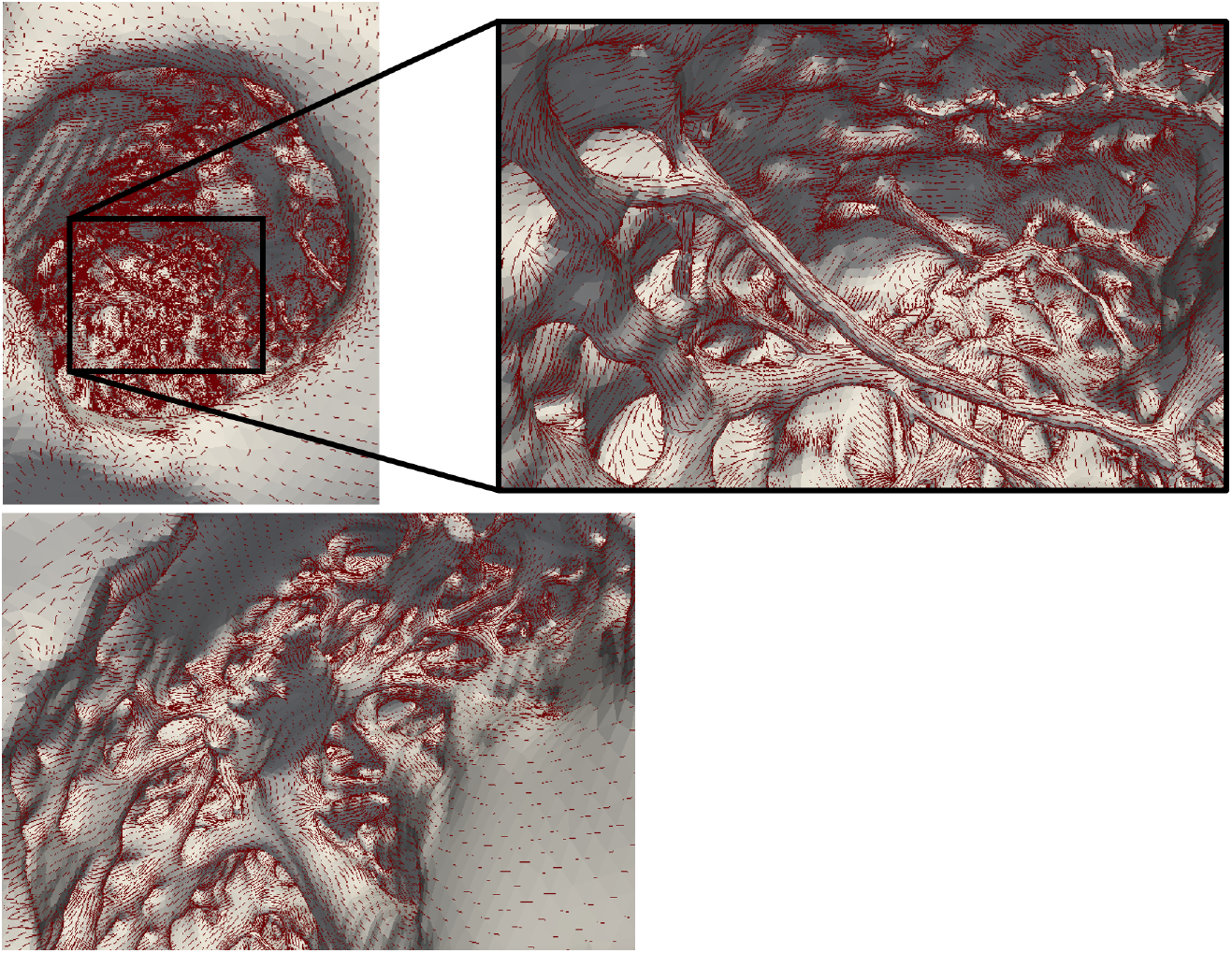
Illustration of the level of detail in the models (geometry **A**) with fibers, represented by red lines, overlayed. **Top:** Left ventricular endocardium with focus on the longest false tendon. **Bottom:** Right ventricular endocardium.

### Activation Locations

The location of the activation points within the biventricular cavities was set following the work of Durrer et al. [18]. The initial activation regions (IARs) are displayed in Fig 3 and labeled as follows: **1-2**. two activation areas on the RV wall, near the insertion of the anterior papillary muscle (A-PM) **3**. high anterior para-septal LV area below the mitral valve **4**. central area on the left surface of the septum **5**. posterior LV para-septal area at 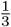 of the apex-base distance. The same IARs location was used for each detailed/smoothed geometrical couple.

**Fig 3.**
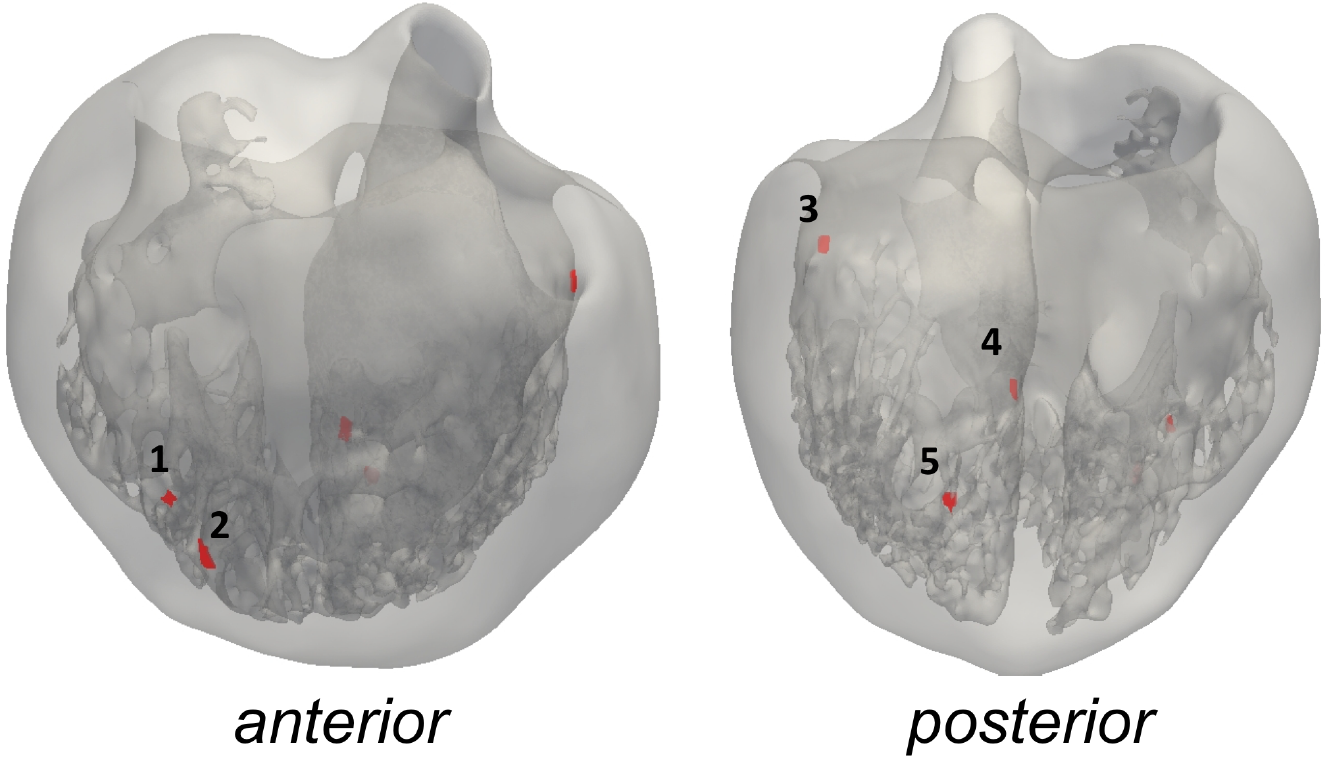
Anterior and posterior view of the geometry **C** with activation points. Points 1-2 are right ventricular IARs, 3-5 – left ventricular IARs).

### Parameterization of Diffusion

As anticipated in Section 2.3, a faster-conducting endocardial surface condition, including both trabecular and FTs network, was used in this work so to simulate the Purkinje fibre network. In order to determine which diffusion to assign to the entire myocardium (*D*_*m*_) and which to apply only on the endocardial surface (*D*_*e*_), thorough analyses were carried out varying both *D*_*m*_ and *D*_*e*_ in all detailed geometries. Diffusion in both myocardium and endocardium were varied, taking as reference the diffusion value in the work from Bueno-Orovio et al. [19] for the compact myocardium; while for the endocardium we employed a higher diffusion to account for the Purkinje network and obtain physiological values of both total activation times (TATs) and conduction velocity (CVs). The selected myocardial and endocardial diffusion values were determined in order to obtain human physiological values of QRS interval and conduction velocities as reported [20] and [21] in all geometries. This objective was important to discard confounding aspects that could arise due to diffusion coefficient differences between models.

#### Myocardial Infarction

A cardiac MRI scan was performed on a patient undergoing electrophysiological studies by our clinical collaborators. Three-dimensional delayed-enhanced acquisitions (1.4×1.4×1.4*mm*^3^) were based on a inversion recovery turbo-field echo sequence (IR-TFE). The entire sequence was triggered with a respiratory navigator to compensate for minor volume displacements. Scar segmentation was performed following the methodology of Lopez-Yunta et al. [22]. Scar registration ensured the transmurality of the scar on the new anatomy. The heterogeneous scar was parameterized following the experimental measurements of [23], with an isotropic diffusion equivalent to the transverse diffusion employed for the normal myocardium. The dense scar was modelled as an inactive material with an extremely small isotropic diffusion equivalent to 10% of the normal extracellular diffusion used in [24], which assumes that some interstitial fluid still exists within the dense scar. The scar was registered to both the smooth and detailed geometries of heart *D* as shown in Figure 8. The clinical protocol for risk stratification of Mark Josephson [25] was employed to assess the ventricular tachycardia risk on the smooth and detailed anatomy with male and female action potential phenotypes. The selected S1 basic cycle length for the programmed stimulation protocol was set to 400 ms. Three stimuli locations were employed: apex of left ventricle (LV Apex), right ventricle (RV Apex) apex, and right ventricular outflow tract (RVOT)

### Data Analysis

To assess the influence of anatomy and gender phenotype on electrophysiology simulations, all heart anatomies were simulated with both male and female phenotypes in order to dissect these gender-specific differences from the anatomical variations.

A pseudo-electrocardiogram (pseudo-ECG) was calculated by locating each biventricular model within a torso and recording the cardiac electrical activity at three field coordinates (or “electrodes”) positioned at the approximate location of the right arm (RA), left arm (LA) and left leg (LL). The pseudo-ECG was calculated as an integral over the spatial gradient of the transmembrane voltage within the cardiac tissue. The pseudo-ECG is calculated without considering the tissue conductivity within the torso between the heart and the leads [26, 27]. A detailed explanation on the calculation of the pseudo-ECG can be found in the supplemental data section S 1.4.

The following quantities of interest were analysed:

***Trabecular volume*** was calculated as the percentage change between the volume of the detailed (*V*_*d*_) and the one of the smoothed (*V*_*s*_) biventricular geometries: 100·(*V*_*d*_ −*V*_*s*_)*/V*_*d*_.

***Total activation time*** (*TAT*) was calculated as the time at which the biventricular geometries were fully depolarized at V ≥ 0mV.

***Conduction velocity*** (*CV*) was calculated as the mode of the conduction velocity distribution on the epicardial surface of the geometries.

***QRS interval duration*** (*QRS*) was calculated as the time at which the QRS on the pseudo-ECG of the three leads (LI, LII and LIII) reached the baseline voltage.

***QT interval duration*** (*QT*) was calculated as the time at which the QT wave on the pseudo-ECG of three leads (LI, LII and LIII) reached the baseline voltage.

***QT interval duration difference*** (*QT*_*diff*_) was calculated as the difference between the QT interval duration between the female (*QT*_*f*_) and male (*QT*_*m*_) phenotype on the same geometry: *QT*_*diff*_ = *QT*_*f*_ − *QT*_*m*_.

***QRS duration difference*** (*QRS*_*diff*_) was calculated as the difference between the QRS duration between the female (*QRS*_*f*_) and male (*QRS*_*m*_) phenotype on the same geometry: *QRS*_*diff*_ = *QRS*_*f*_ − *QRS*_*m*_

The activation pattern of the detailed models was qualitatively analyzed and compared to the smoothed cases.

***Sustained Ventricular Tachycardia*** was characterised by three or more reentrant full heart activations during the S1-S2-S3-S4 pacing protocol. Any other arrhythmic events are defined as reentries that do not sustain for more than 2 beats.

## Results

### Diffusion Analyses

After conducting a thorough study on all four biventricular geometries, a single set of diffusion coefficients for the fast-conducting endocardium and myocardial tissue were identified for all hearts. The objective was to find an approximate normal value of diffusion that can reproduce normal human electrophysiological behaviour in various detailed anatomies. The values obtained for the myocardium were 5.8 · 10^−3^ *cm*^2^*/s* in the fiber direction and 1.9 · 10^−3^ *cm*^2^*/s* in the transverse and normal directions. For the purkinje layer, the diffusions obtained were 1.7 · 10^−2^ *cm*^2^*/s* in the fiber direction and 5.8 · 10^−3^ *cm*^2^*/s* in the transverse and normal directions.

### Electrophysiology Simulations

Results from electrophysiological simulations of the four detailed and smoothed biventricular models with both male and female phenotype are listed in Table 2 and Table 3 respectively. As can be seen, female phenotype prolonged the QT interval in all cases. A two-tailed unequal variance t-test (Welch t-test) was used to compare groups of measurements and the null hypothesis was rejected at 0.05. Noticeable local differences on the activation pattern between the detailed and smoothed geometries can be observed in Fig 4. Local distribution of the depolarization wave is shown at 60 ms for both detailed and smoothed cases with male phenotype. False tendons provide a fast activation bridge that change the activation pattern in the detailed geometries as observed in Fig 5.

**Table 2.**
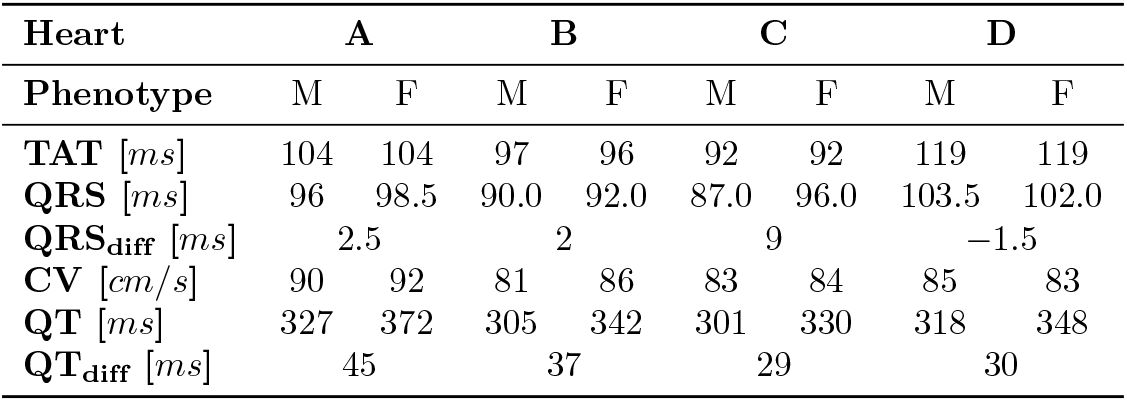
Electrophysiological simulation results, detailed geometries.

**Table 3.** Electrophysiological simulation results, smoothed geometries.

| Heart | A |  | B |  | C |  | D |  |
| --- | --- | --- | --- | --- | --- | --- | --- | --- |
| Phenotype | M | F | M | F | M | F | M | F |
| **TAT** [ms] | 100 | 100 | 84 | 84 | 94 | 94 | 120 | 120 |
| **QRS** [ms] | 98.0 | 104.0 | 81.0 | 83.5 | 90.5 | 91 | 97.5 | 103.5 |
| **QRS<sub>diff</sub>** [ms] | 6 |  | 2.5 |  | 0.5 |  | 6 |  |
| **CV** [cm/s] | 80 | 80 | 88 | 86 | 85 | 86 | 86 | 86 |
| **QT** [ms] | 341 | 373 | 305 | 335 | 300 | 334 | 318 | 349 |
| **QT<sub>diff</sub>** [ms] | 32 |  | 30 |  | 34 |  | 31 |  |

**Fig 4.**
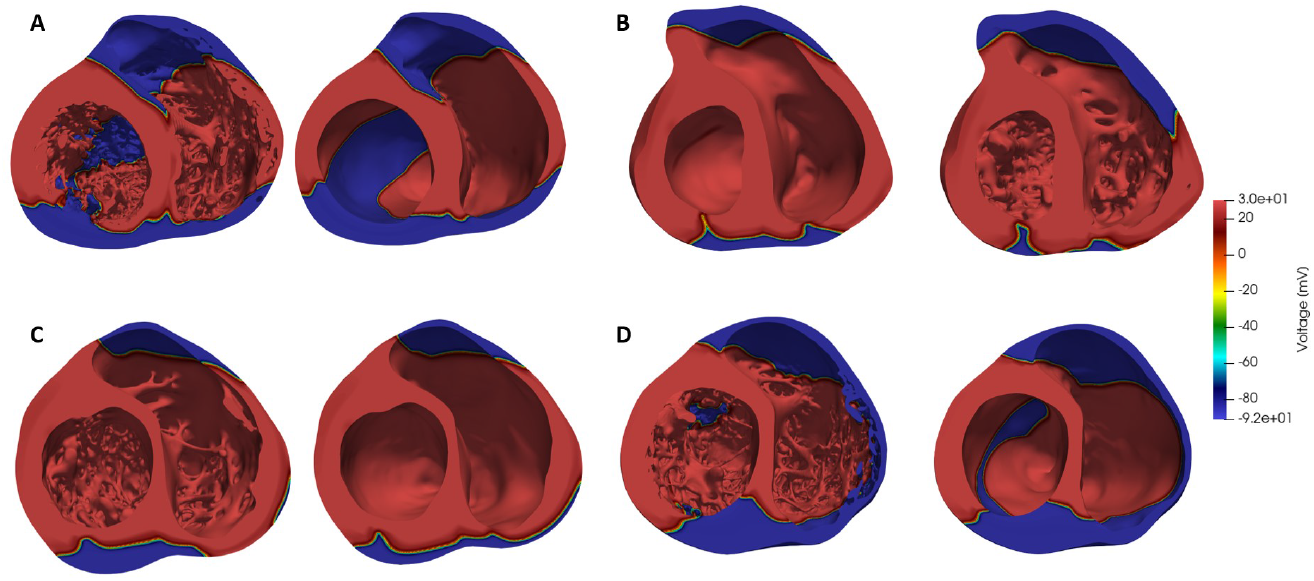
Depolarization wave distribution at 60 ms, male phenotype.

**Fig 5.**
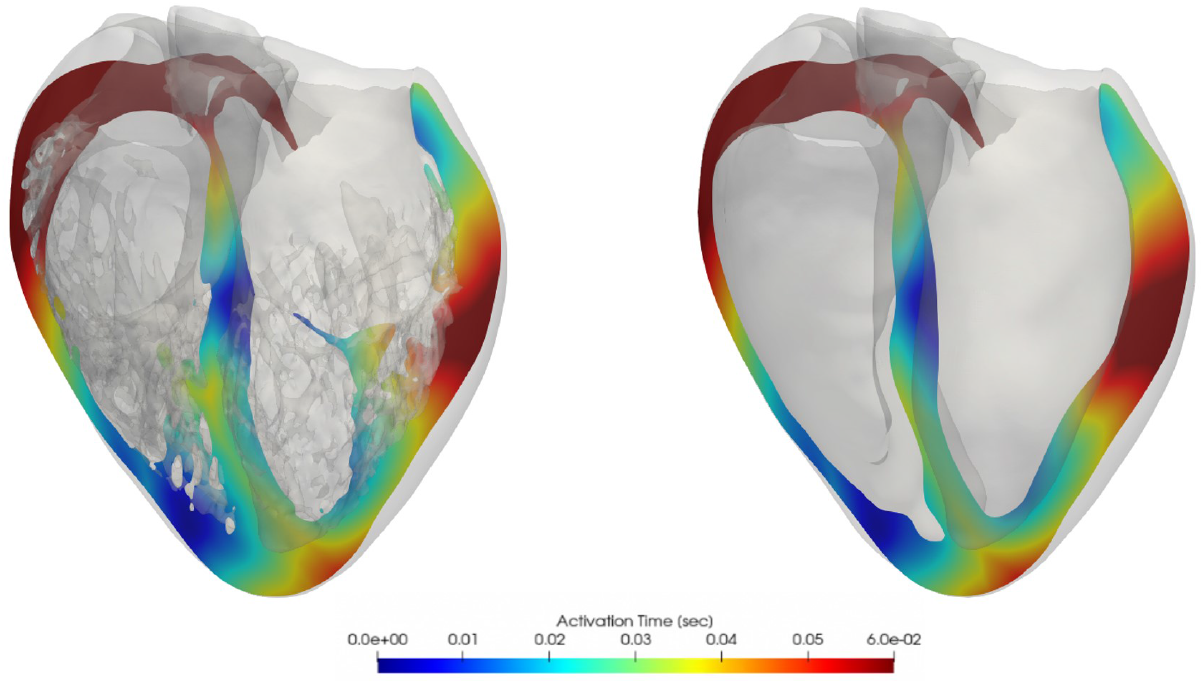
Cross-sectional slices of ventricular isochrone activation maps, smoothed (left) and detailed (right) heart **D** comparison, male phenotype.

### Pseudo-ECGs

Fig 6 shows the obtained pseudo-ECG for Lead I, II and III, for smoothed and detailed male (**A-B**) and female (**C-D**) geometries respectively, with both phenotypes. QT prolongation is observed in all female phenotypes cases, as expected. The root-mean-squared difference (RMS) between the pseudo-ECG of the smoothed and detailed geometries is reported, for both male and female phenotypes, in Table 4. It is noticeable that the differences between the smoothed and detailed geometries is more pronounced on the QRS segment morphology. The T wave is, however, the most different feature in the pseudo-ECGs between genders. Interestingly, heart C, which has the smallest trabecular volume, also has the smallest difference between the detailed and smoothed pseudo-ECG RMS error.

**Table 4.** Root mean square difference between the pseudo-ecg waveform of the detailed- and smoothed-geometry simulations on both female and male gender phenotypes.

| Heart | Phenotype | Lead I (mV) | Lead II (mV) | Lead III (mV) |
| --- | --- | --- | --- | --- |
| A | Female | 2.2 | 3.8 | 4.9 |
| B | Female | 2.1 | 3.1 | 2.1 |
| C | Female | 0.8 | 1.9 | 2.0 |
| D | Female | 1.8 | 3.9 | 3.7 |
| A | Male | 2.3 | 3.8 | 4.8 |
| B | Male | 2.3 | 3.1 | 2.2 |
| C | Male | 0.9 | 1.9 | 2.1 |
| D | Male | 1.5 | 4.2 | 3.9 |

**Fig 6.**
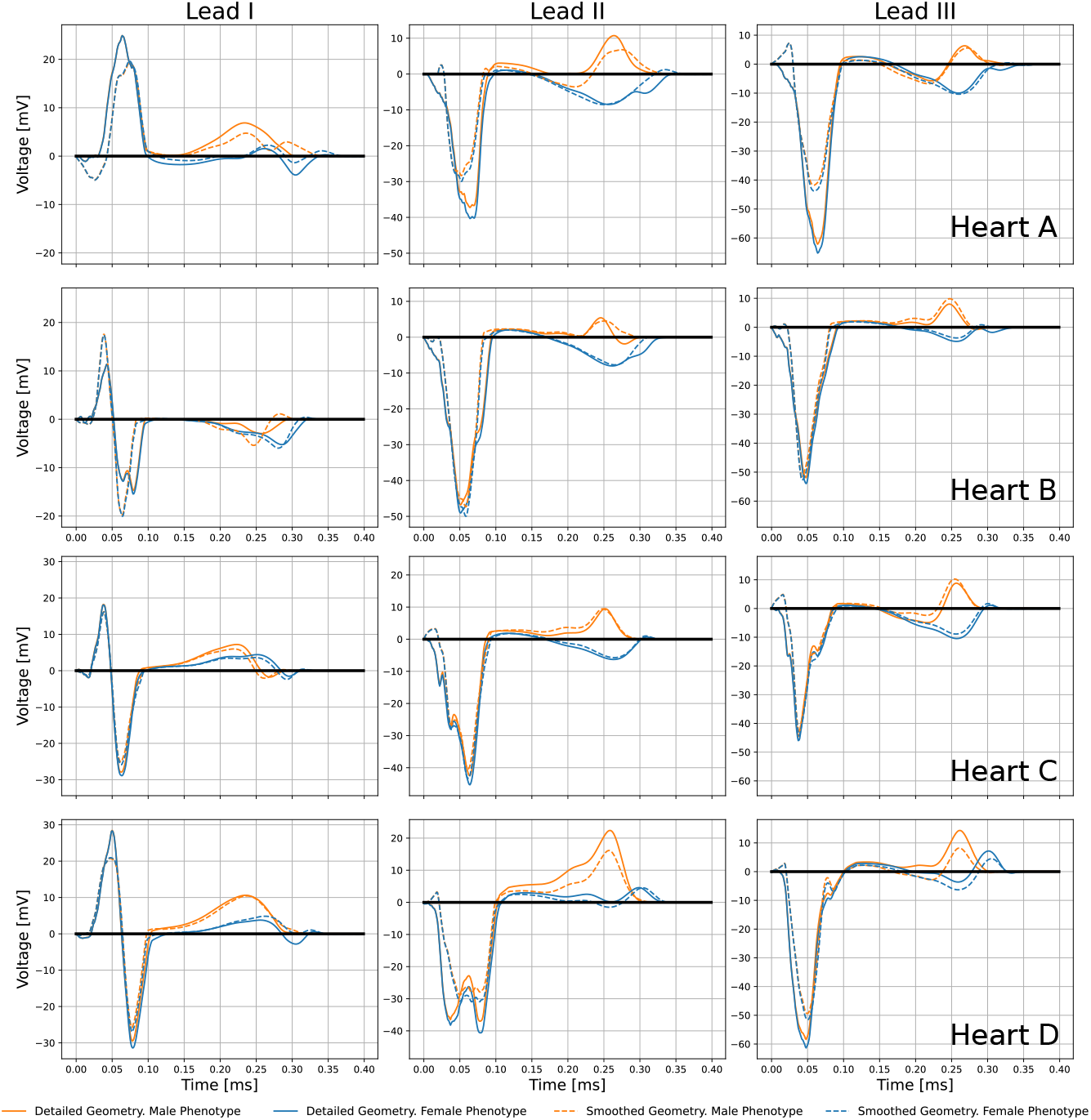
Lead I, II and III pseudo-ECG results for smoothed/detailed male (**A-B**) and female (**C-D**) geometries with both male/female phenotypes.

### Ventricular Tachycadia Inducibility

A total of 126 simulations were done to reproduce the programmed stimulation protocol commonly performed in the clinical setting (S1-S2-S3-S4) to stratify the risk of postinfaction cardiac arrest. The arrhythmic events throughout the inducibility study are detailed in Table 5. The results show a high number of arrhythmic events (48%) in the male phenotype, detailed heart, while the smooth geometry of the same gender phenotype shows a 14% of events. For the female phenotypes of the detailed and smoothed geometries only a 6% and 1% of arrhythmic events, respectively, occurred throughout the entire protocol. The female phenotype simulations elicited a sustained VT with an RV apex stimulus on both, the detailed and smooth geometries. Ventricular tachycardia occurred only with an LV apex stimulus in the case of the detailed, male geometry while sustained ventricular tachycardia occurred on both RV Apex and RVOT stimulus locations in the case of the smooth male geometry.

**Table 5.** Arrhythmic events including reentries and sustained VTs. The number of events (E) are shown along with the total number of simulations (S) for each programmed stimulus protocol at each location.

| Stimulus Location | RV Apex | RVOT | LV Apex | Total |
| --- | --- | --- | --- | --- |
| Events/Simulations | E/S | E/S | E/S | E/S |
| Detailed Female | 2/13 | 0/12 | 0/8 | 2/33 |
| Smooth Female | 1/11 | 2/10 | 0/8 | 3/29 |
| Detailed Male | 4/10 | 7/15 | 6/10 | 17/35 |
| Smooth Male | 2/10 | 1/9 | 1/10 | 4/29 |

Each VT was analyzed in order to locate the channels that enabled the tachycardia to sustain. Fig 7 and 8 show the comparisons between geometries and phenotypes for each simulation with a sustained VT. The VT on both female phenotype geometries occurred at an S1=400, S2=300, S3=300 and S4=300. Interestingly, on both geometries the initial reentry location is exactly the same, and it is marked as a blue sphere in Fig 7. The channel locations for the subsequent reentries enabling the VT are completely different, and marked with red spheres. The two female VT simulations provide different reentry cycle lengths. The detailed geometry presents reentries of variable timings: 363, 420, 410, 420, 380, 350 and 370 ms and all are located at the posterior epicardial wall. The smoothed geometry reentrant cycle lengths were 300, 700 and 400 ms and only one of the reentry channels was located at the epicardium.

**Fig 7.**
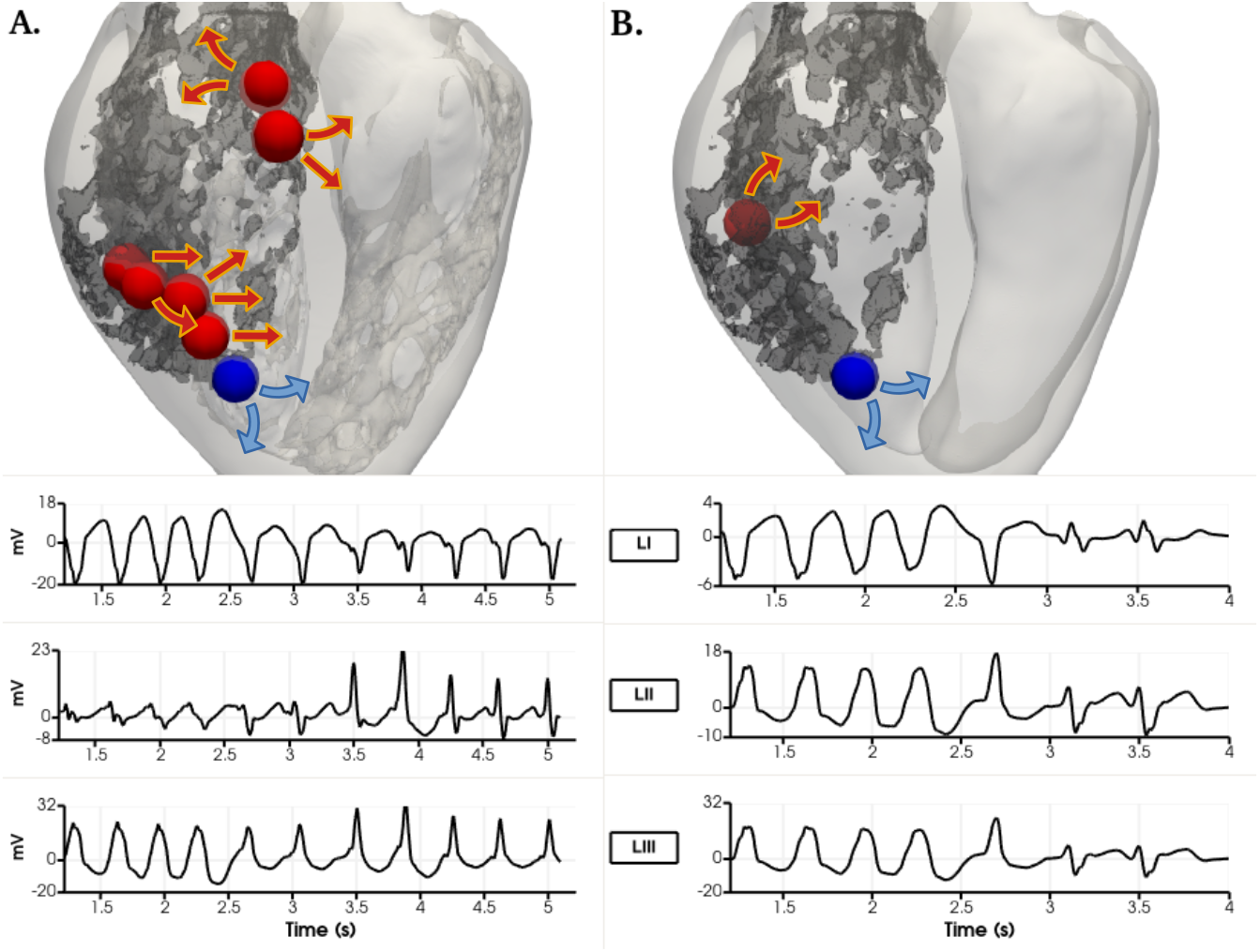
Ventricular tachycardia reentry channels in the female phenotype simulations with right ventricular apex stimulus location at S1=400, S2=300, S3=300, S4=300 ms. View from the epicardial posterior side of the biventricular geometry. Dense scar marked in dark grey. Arrows show the propagation wavefront of each reentry. A. Detailed geometry. B. Smoothed geometry. Their corresponding Pseudo-ecg is shown below for the LI, LII and LIII leads. The sphere in blue shows the exact same reentry channel on detailed and smoothed simulations.

**Fig 8.**
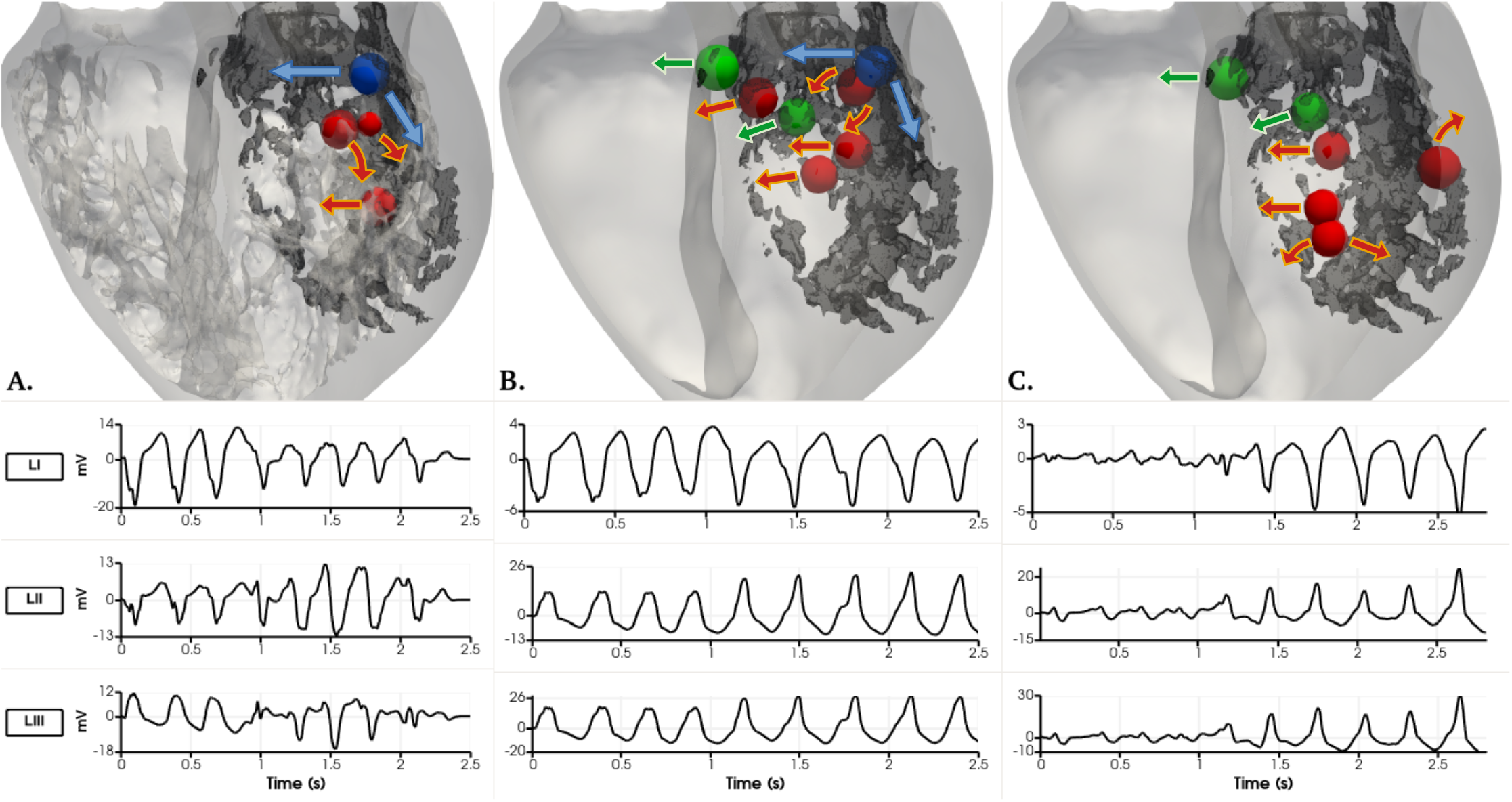
Detailed and smoothed male VT simulations. The dense scar is seen in dark grey colour on all three geometries for reference. View from the mid longitudinal plane towards the posterior side of the heart. Arrows show the propagation wavefront of each reentry. A) Four reentry channel locations on the detailed male geometry with an LV apex stimulation shown as spheres. B) Seven reentry channel locations on the smoothed male geometry with an RV apex stimulation shown as spheres. The blue sphere on A and B corresponds to the same location. C) Six reentry channel locations on the smoothed male geometry with a RVOT stimulus shown as spheres. The green spheres on B and C correspond to the same channel location. Underneath each geometry, the three pseudo-ECG leads recorded for each case.

Sustained VT was obtained on the detailed, male phenotype simulations at S1= 400, S2=270, S3=260, S4=240 ms using the LV apex stimulus location. The tachycardia is of a more irregular nature with basic cycle lengths of 270, 270 and 290 ms as observed in Fig 8. The reentry channel for the initial three re-entrant beats is located at the LV posterior wall, approximately in the center of the scar. Interestingly, a trabeculae (small, red sphere in Fig in 8A.) created a pathway to create a reentry channel, which created a fast pathway of re-activation towards the apical septum region through the endocardial structures. The last reentry channel is located towards the base of the posterior LV wall, shown in blue.

The sustained tachycardia obtained in the smoothed, male geometry occurred during RV apex stimulation at S1= 400, S2=260, S3=250, S4=240 ms. Seven reentry channels were identified during a 2.8 second simulation. Two are located at the septum, two more channels on the posterior LV wall. The VT ultimately sustains through a channel at the basal location shown in blue in Fig 8. Interestingly the two reentry channel basal locations shown in blue on the detailed (A.) and smooth (B.) anatomies in Fig 8 are the only channels that correspond between the two simulations. In the case of the smoothed, male geometry VT generated using the RVOT stimulation location at S1= 400, S2=260, S3=230, S4=240 ms, six channels are located to create the reentrant waves: on the basal section of the septum, the basal section of the LV free wall, and various reentry beats arising at the posterior LV wall, approximately below the papillary muscle. Interestingly, only two channels, after an RV apex and RVOT stimuli on the smooth male geometries shown in green in the Fig 8 B. and C. coincide. All pseudo-ecg recordings are provided for detailed analysis. All of the male phenotype simulations elicited reentries at the mid-myocardial or endocardial transmural regions. A video of each VT simulation can be found in the supplemental data.

## Discussion

The cardiac ventricular endocardium of humans are highly trabeculated and complex, such anatomical characteristics are often ignored in ventricular electrophysiology studies. This is the first time, to the best of our knowledge, that all small endocardial sub-structures with cross-sectional area ≥1 mm2, were utilized in human cardiac electrophysiology simulations; here we modeled four detailed, biventricular, anatomically normal human hearts. Until now, gender phenotype has been another characteristic often neglected, which was also considered in our study. It is important to note that all bio-markers were analyzed using statistical tools even though the sample number is small (four to eight samples). Obtaining high resolution ex-vivo imaging data of normal human hearts is extremely difficult, and therefore only four anatomies were employed. Yet, the authors also understand there are limitations to the significance of these results, but the analyses of an even larger data set is an active part of our ongoing work.

### Diffusion Parameterization

The objectives for performing thorough diffusion coefficient analyses were to obtain the approximate values that would produce physiological values of TATs and CVs on all the detailed geometries. The selected diffusion coefficients were employed to establish the baseline condition for all our human heart simulations. This was specifically done to eliminate any confounder results due to different diffusion coefficients among hearts. It is however, important to note that the optimal diffusion coefficient estimated corresponded to our assumption of a fast-endocardial diffusion: i.e., that was needed to compensate for the fact that we did not include Purkinje networks. When no fast-endocardial condition was included, it was impossible to obtain normal TATs and CVs (within measured ranges) in all of our human heart simulations.

### Gender differences in detailed and smoothed geometries

The two-sample Welch t-test provided significant differences on the QTs relative to gender; i.e., between both male and female detailed and smoothed geometries (p = 0.02 and p = 0.048, respectively). It was expected that the female phenotypes would present longer QT intervals as expected from literature [3]: the average QTs prolongation was 35.25 ms on the detailed geometries and 31.75 ms on the smoothed ones.

We hypothesized that including gender-specific phenotypes would be important when simulating cardiac arrhythmia, since QT-interval values may influence the triggering and/or sustaining of arrhythmias; i.e., particularly for reproducing the cycle length of a given patient-specific arrhythmia. Additionally, statistically significant longer QRS durations were found for female phenotypes (p=0.013), regardless of geometry. Average values for the female QRS duration was 96 ms versus 92.9 ms for the male. Finally, no gender statistically significant differences were observed for either TATs and CVs.

### Differences in pseudo-ECG

The pseudo-ECG calculations presented here were not intended to be compared to physiological waveforms, given the limitations of our simulations: the relative volumes of the torso were not solved using a forward solution and the same torso anatomy was used to provide the lead coordinates for all four hearts; furthermore, we employed a monodomain approach for conduction propagation. Again note, that small variations in rotations or translations could impact such lead information. In addition, we had no functional data for these particular subjects and the activation sequences employed were generalization from the data measured by [18]. Our main uses of the pseudo-ECGs were to find the variations between the anatomies and phenotypes. Results show that QRS complexes followed almost the same profiles for both male and female phenotypes, but were different when compared between smoothed and detailed ventricular geometries. The predicted differences of the QRS complexes between detailed and smoothed geometries correspond to the different regional activation patterns observed in the absence of endocardial structures. Furthermore, it is also noticeable that on some leads, the QRS wave appears fractionated in the detailed anatomies, feature which disappears in the smoothed anatomies. The T waves showed clear differences, as can be observed from Figure 6; the female phenotypes introduced a prolongation of the QT interval (in both smoothed and detailed geometries), behavior which is linked to the higher repolarization time in the female action potential. Interestingly, the T wave of Lead I was inverted on the child heart, case **B** given the same activation conditions than the adult heart simulations. It is plausible that slight rotations within the torso can be the reason for the T wave inversion in this case, or the fact that the torso was that of an adult size. Also noticeable is the T-wave inversion in the female phenotype, mainly on Leads II and III. There was also a slight reduction of the T-wave magnitude within the smoothed geometries in comparison to the detailed cases, particularly in hearts **A** and **D**. The smoothed geometries introduced an average RMS error on each pseudo-ecg lead of approximately 1.73, 3.2 and 3.2 mV for both male and female phenotypes (see Table 4). The measured errors between the smoothed and detailed anatomies seem to be directly correlated to the trabecular volume in each heart.

It is clear that gender played an important role on the pseudo-ECG morphologies, particularly on the T waves. Hence we can conclude that the absences of endocardial structures modifies the QRS complexes and/or the T-waves in complex manners; i.e., depending on the subject-specific trabecular structures and the activation and repolarization sequences on any computed lead.

### Differences between detailed and smoothed geometries

No significant QT-interval differences were found between detailed and smoothed geometries, for either the male and female phenotypes (p= 0.78 and p= 0.99 respectively). Anatomical differences produced QRS variations that do not correlate to the trabecular volume of each case. In some cases the QRS duration increased between detailed and smooth versions, while on others it decreased. The average of the absolute value differences was 5.1 ms on both male and female phenotypes. The largest QRS variations occurred on heart **B**, where the smooth geometry decreased the QRS duration in both male and female phenotypes. In the other hand, heart **A** showed an opposite behaviour, where QRS duration was longer in the smoothed geometry for both male and female phenotypes. Heart anatomies **C** and **D** had gender-dependent QRS change variations between detailed and smooth geometries.

Moreover, larger regional differences in activation times, caused by overall different propagation paths (lack of shortcuts) of the depolarization waves, were found in the smoothed cases, although there were no consistent effects on global TATs. As one can observe in Figure 4, indeed, smoothed geometries were characterized by different propagation paths of the depolarization waves as compared to the detailed anatomic cases. This was due to the fact that the presence of FTs and trabeculae provide propagation shortcuts and thus helped the depolarization waves to spread across distal endocardial regions.

TATs (p= 0.60) and CVs (p= 0.65) did not present any significant differences between the employed detailed and smooth geometries. However, it is noticeable that heart **A** and **B** showed longer TAT in the detailed anatomy in comparison to their smooth counterpart for both genders. Finally, using as example the human heart model **D**, the differences on the local depolarization could be appreciated. On the one hand, the position of a long FT: i.e., crossing the LV cavity and attaching to the septum near one of Durrer’s activation locations, thus providing a fast shortcut of the depolarization wave. The impact of the main FTs on ventricular activations were clearly observable in Figure 5, where cross-sectional slices representing the activation times at the level of the main FT are shown. As can be observed, the presence of a long FT within the LV, provided a clear propagation shortcut for the depolarization wave.

### Differences between gender phenotypes and detailed and smoothed geometries regarding the inducibilities of tachycardia

The inducibility protocol employed here provided evidence of the relative impacts of endocardial structures as well as the gender phenotypes on our computational simulation results. Utilization of our described set of human hearts, with the same myocardial infarction registered onto each, provided different risks for arrhythmic events relative to genders. Furthermore, the reentry channels identified in the same cardiac geometry, and relative to gender phenotypes were both different. When we compare the reentry channel locations within the detailed versus the smoothed geometries, it was clear that they were more often than not different; only in some occasions did they coincide. It was highly interesting from our perspective, that the induced VTs occurred at different stimuli locations between the simulated smooth and detailed geometries, and even more important was that the refractory periods also differ according to the employed anatomies and/or locations of the stimuli. The results regarding the inducibility of tachycardia may provide physiologically relevant information regarding the risk of females to VT and its recurrence at different hormonal stages during their life time. It is highly interesting that while the male phenotype produced a much higher percentage of arrhythmic events throughout the programmed stimulation protocol than the female phenotype, the VT induced in the female detailed anatomy proved to be sustained for a much longer time. Another physiologically relevant aspect of this work is that it may provide evidence of why a VT morphology may change at different hormonal stages. Furthermore, it provides evidence of the importance of the personalization of the approximate repolarization time on patient-specific VT simulations.

## Limitations

One of the primary limitations of the present study was the somewhat small number of human hearts analyzed in this study. Yet, it may also be possible that with analyses on a larger subject sampling, our findings regarding both the diffusion coefficient analyses and gender/anatomical differences could potentially provide more statistically significant results. Nevertheless, it is important to also consider that collecting such normal human data with a high level of anatomical details was not a trivial undertaking and much effort has been performed to present the current computational simulations.

A second limitation that should be noted was in the current analyses the Purkinje systems were not included in these models. We used instead a fast-endocardial condition and we provided sensitivity analyses to estimate the optimal myocardial and endocardial diffusion coefficients. Although it is possible to extract and represent computationally the free-running Purkinje system in our geometries, this would also represent an approximation far from histologically measured Purkinje networks running within human ventricles. The optimal way to validate the simulations in terms of the activation systems would be to image the Purkinje system from high-resolution contrast enhanced micro-CT of human hearts, as what was done by [28]. Work is ongoing to do so, but this may take years to accomplish.

The fact that all four anatomies were artificially, spatially located within a generic single torso geometry, makes it impossible to directly correlate the lead information to clinical electrocardiographic recordings. The pseudo-ECG recordings presented cannot be clinically validated.

Finally, only cardiac electrophysiologic quantities of interest were analyzed in this current study; i.e., there was no consideration relative to the effects of electro-mechanical coupling. In other words, the lack of mechano-electric feedback in our described simulation, may provide different results versus fully coupled electro-mechanical simulations. Nonetheless the results presented in this paper provide novel insights relative to the effects of gender phenotypes as well as and the roles of trabeculae and FTs on the overall cardiac electrophysiologic behavior.

### Conclusion

Electrophysiological simulations of four human biventricular heart biventricular models were presented with high geometrical details, which to the best of our knowledge, has not been reported yet in the electrophysiological simulation literature. This study uniquely represents some of the first insights in human cardiac electrophysiological simulations relative to assessing the roles of both endocardial substructures and gender phenotypes. This work provides strong evidence that neglecting either endocardial anatomical details or cell-level gender characteristics for cardiac electrophysiology simulations may lead to erroneous results. Our work indentified a significant QT-interval increase in the detailed female human heart phenotypes; i.e., shown also by the pseudo-ECGs results, coinciding with the reported longer QT prolongations in female hearts. QRS interval durations were also significantly longer in the female phenotypes. Therefore the pseudo-ECG patterns are subject to variations due to both anatomy and gender. Additionally, our simulations suggest that the absence of trabeculations introduces regional differences in the propagation pathways relative to the depolarization wavefront. The presence of FTs were considered to shortcut the signal propagation leading to differences in local activation patterns. Finally, it was shown that the levels of endocardial details and differences in the gender phenotypes also will likely affect the arrhythmic risks and conduction channel locations: i.e., during the in-silico, clinical assessments of tachycardia inducibilities.

Our future work includes the analyses of larger datasets, in order to verify statistical significances, in addition to including the effects of the mechano-electric feedback on clinically relevant pathological scenarios, thus both aiming to increase the accuracy of cardiac computational model predictions for their clinical translation.

## Data Availability

The data table that contains the markers processed to analyse the results from the simulations are all included within the manuscript. The anatomically detailed human heart images can be found within the Visible Heart Laboratory database (http://www.vhlab.umn.edu/atlas/histories/histories.shtml). Volumetric transmembrane potential data is unavailable due to its large storage requirements. Videos of the ventricular tachycardias produced are available as supplementary data.

## Supporting Informatio

**Anatomical mesh construction**

**Electrophysiology solver**

**Gender phenotypes**

**Pseudo-ECG calculation**

**Video. Ventricular tachychardia media**

## Acknowledgments

The DICOM datasets were provided by the Visible Heart® Laboratories, obtained by MRI scanning of perfusion fixed hearts that were graciously donated by the organ donors and their families through LifeSource.

Part of the simulation computing hours were provided by the CompBioMed project and the Red Española de Supercomputación (RES) (Activity IDs: FI-2018-2-0049 and BCV-2019-2-0014).

The clinical images of the myocardial infarction employed were provided by Dr. David Filgueiras-Rama, from Hospital Clinico San Carlos and Fundación Centro Nacional de Investigaciones Cardiovasculares.

This project has received funding from CompBioMed from the European Union’s Horizon 2020 research and innovation programme under grant agreement No 675451 (phase 1) and grant agreement No. 823712 (phase 2). This project has also received funding from SilicoFCM, from the European Union’s Horizon 2020 research and innovation programme under grant agreement No 777204, and was partially funded by Spanish NEOTEC Programme EXP-00123159/SNEO-20191113.

JA-S is funded by a Ramon y Cajal fellowship (RYC-2017-22532), Ministerio de Ciencia e Innovación, Spain. JA-S is also funded by Plan Estatal de Investigación Científica y Técnica y de Innovación 2017-2020 from the Ministerio de Ciencia e Innovación y Universidades (PID2019-104356RB-C41/AEI/10.13039/501100011033): meHeart ME PID2019-104356RB-C44.

CB is funded by the Torres Quevedo Program (PTQ2018-010290), Ministerio de Ciencia e Innovación, Spain.

LKGEM was funded by Fundacion Carolina-BBVA.

