## Supplemental data for "Ventricular anatomical complexity and gender differences impact predictions from computational models"

### S1 Appendix.

#### S1.1 Anatomical Mesh Construction.

The software Fiji[1] [1] was used to carry out high resolution MRI segmentation, with the maximum entropy-based thresholding algorithm [2]. The images were characterized by excellent contrasting gray scales, which permitted thresholding to achieve biventricular heart models with highly detailed endocardiums (endocardial structures included were $\geq 1$ mm$^2$ in cross-section). The reconstruction of four biventricular meshes including detailed endocardial structures was accomplished utilizing Seg3D [3] marching cubes algorithm. The obtained surface meshes were uniformly remeshed with the platform Remesh [4] and volumetric tetrahedral meshes were generated using ANSYS ICEM CFD (ANSYS® Academic Research Mechanical, USA [5]). Wireframe images showing the element distribution within each heart anatomy are shown in 1.

#### S1.2 Electrophysiology solver.

Finite Element Method (FEM) and Finite Difference Method (FDM) were used for the space and time discretisation of the electrical activation potential propagation equation in (**??**). Once discretized, the equation becomes:

$$\left(\frac{M}{\Delta t} + \theta G\right) \Delta\phi + M^{\mathrm{d}} I_{\mathrm{ion}} = -G\phi^{\mathrm{n}} \qquad (1)$$

where $\Delta\phi = \phi^{\mathrm{n+1}} - \phi^{\mathrm{n}}$ is the unknown difference between two time steps, M is the mass matrix, G is the electrophysiological stiffness matrix, and $\theta$ determines whether the time integration scheme is first order explicit, Forward Euler ($\theta = 0$), first order implicit, Backward Euler ($\theta = 1$) or second order implicit, Crank-Nicholson ($\theta = 0.5$).

[1] https://imagej.net/Fiji

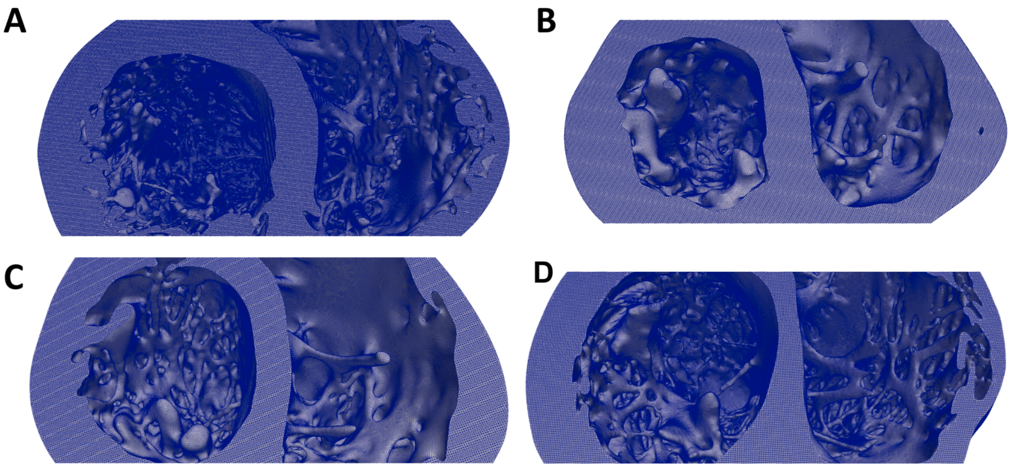


**Fig 1.** Male (**A-B**) and female (**C-D**) anatomically detailed biventricular wire-frame meshes, mid-cavity sections. Elements are tetrahedral and regularly sized throughout the whole anatomy. Element size is the same in all four anatomies.

The discrete equation is then solved using a first order Yanenko operator splitting as follows:

$$Cell\ model(\Delta\phi^*) : \frac{\Delta\phi^*}{\Delta t} + I_{\mathrm{ion}}(\phi) = 0 \tag{2}$$

$$Tissue\ model(\Delta\widetilde{\phi}) : \left(\frac{M}{\Delta t} + G\right)\Delta\widetilde{\phi} = -G\phi^* \tag{3}$$

$$Update(\phi^{\mathrm{n+1}}) : \phi^{\mathrm{n+1}} = \phi^{\mathrm{n}} + \Delta\phi^* + \Delta\widetilde{\phi} \tag{4}$$

where the *Cell model* is solved explicitly using a Forward Euler scheme and the *Tissue model* is solved implicitly with either a Backward Euler or a Crank-Nicholson scheme s described by [6].

#### S1.3 Gender phenotypes.

**Table 1. Ion channel conductances ratios relative to male endocardial cell.** No change on mid-myocardial cells was applied.

| Ion channel | Endocardium Male | Endocardium Female | Epicardium Male | Epicardium Female |
|---|---|---|---|---|
| $I_{Ks}$ | 1 | 0.83 | 1.04 | 0.87 |
| $I_{Kr}$ | 1 | 0.79 | 1.09 | 0.875 |
| $I_{K1}$ | 1 | 0.86 | 0.98 | 0.74 |
| $I_{to,s}$ | 1 | 0.64 | 0.6 | 0.26 |
| $I_{NaK}$ | 1 | 0.79 | 0.94 | 0.7 |
| $I_{pCa}$ | 1 | 1.6 | 0.88 | 1.6 |
| $I_{up}$ | 1 | 1.15 | 1.42 | 1.97 |
| $Calm$ | 1 | 1.21 | 1.01 | 1.41 |

#### S1.4 Pseudo-ECG calculation.

The calculation of the pseudo-ECGs derived from the calculation of the unipolar potentials ($\phi_e$) as follows:

$$\begin{aligned}\phi_{\mathrm{e}}(x',y',z') = D\int[-\nabla V_{\mathrm{m}}\cdot(\nabla\tfrac{1}{r})]dx + D\int[-\nabla V_{\mathrm{m}}\cdot(\nabla\tfrac{1}{r})]dy \\ +D\int[-\nabla V_{\mathrm{m}}\cdot(\nabla\tfrac{1}{r})]dz\end{aligned} \tag{5}$$

$$r = [(x-x')^2 + (y-y')^2 + (z-z')^2]^{\frac{1}{2}} \tag{6}$$

where $D$ is the diffusion tensor at every Gauss point, $\nabla V_m$ is the spatial gradient of the transmembrane potential and $r$ is the distance from a source point ($x$,$y$,$z$), which represents a point on the heart geometry, and a field point ($x'$,$y'$,$z'$), which represents the position of one of the electrodes used to calculate the pseudo-ECG (LA, RA, LL). Electrical potential difference was defined as the difference in electric potential between two electrodes; these potential differences are represented as "leads". There is always one exploring (positive) and one recording (negative) electrode. In this way, a propagation wave going towards the exploring electrode produces a positive wave and vice versa. The three leads were defined as:

$$\begin{aligned}Lead_I = \phi_{LA} - \phi_{RA}, \\ Lead_{II} = \phi_{LL} - \phi_{RA}, \\ Lead_{III} = \phi_{LL} - \phi_{LA}\end{aligned} \tag{7}$$

#### S3 Video. Ventricular Tachycardia Media.

Video S1: Ventricular Tachycardia generated on a female phenotype simulation after RV Apex S1-S4 programmed stimulation protocol. Anatomical data was obtained from high resolution MRI ex-vivo human hearts.
Video S2: Ventricular Tachycardia generated on a female phenotype simulation on a smoothed geometry after RV Apex S1-S4 programmed stimulation protocol.
Video S3: Ventricular Tachycardia generated on a male phenotype simulation on a detailed geometry after LV Apex S1-S4 programmed stimulation protocol.
Video S4: Ventricular Tachycardia generated on a male phenotype simulation on a smoothed geometry after RV Apex S1-S4 programmed stimulation protocol.
Video S5: Ventricular Tachycardia generated on a male phenotype simulation on a smoothed geometry after RVOT Apex S1-S4 programmed stimulation protocol.
